# Cardiac Magnetic Resonance Imaging in the German National Cohort: Automated Segmentation of Short-Axis Cine Images and Post-Processing Quality Control

**DOI:** 10.1101/2025.05.20.25328013

**Authors:** Peter M. Full, Robin T. Schirrmeister, Manuel Hein, Maximilian F. Russe, Marco Reisert, Clemens Ammann, Karin Halina Greiser, Thoralf Niendorf, Tobias Pischon, Jeanette Schulz-Menger, Klaus H. Maier-Hein, Fabian Bamberg, Susanne Rospleszcz, Christopher L. Schlett, Christopher Schuppert

## Abstract

**Purpose:** To develop a segmentation and quality control pipeline for short-axis cardiac magnetic resonance (CMR) cine images from the prospective, multi-center German National Cohort (NAKO).

**Materials and Methods:** A deep learning model for semantic segmentation, based on the nnU-Net architecture, was applied to full-cycle short-axis cine images from 29,908 baseline participants. The primary objective was to determine data on structure and function for both ventricles (LV, RV), including end diastolic volumes (EDV), end systolic volumes (ESV), and LV myocardial mass. Quality control measures included a visual assessment of outliers in morphofunctional parameters, inter- and intra-ventricular phase differences, and LV time-volume curves (TVC). These were adjudicated using a five-point rating scale, ranging from five (excellent) to one (non-diagnostic), with ratings of three or lower subject to exclusion. The predictive value of outlier criteria for inclusion and exclusion was analyzed using receiver operating characteristics.

**Results:** The segmentation model generated complete data for 29,609 participants (incomplete in 1.0%) and 5,082 cases (17.0 %) were visually assessed. Quality assurance yielded a sample of 26,899 participants with excellent or good quality (89.9%; exclusion of 1,875 participants due to image quality issues and 835 cases due to segmentation quality issues). TVC was the strongest single discriminator between included and excluded participants (AUC: 0.684). Of the two-category combinations, the pairing of TVC and phases provided the greatest improvement over TVC alone (AUC difference: 0.044; p<0.001). The best performance was observed when all three categories were combined (AUC: 0.748). Extending the quality-controlled sample to include acceptable quality ratings, a total of 28,413 (95.0%) participants were available.

**Conclusion:** The implemented pipeline facilitated the automated segmentation of an extensive CMR dataset, integrating quality control measures. This methodology ensures that ensuing quantitative analyses are conducted with a diminished risk of bias.

## Introduction

Cardiac magnetic resonance imaging (CMR) is widely acknowledged as the standard for noninvasive evaluation of cardiac structure and function. Its ability to provide detailed quantification of ventricular volumes, myocardial mass, and functional parameters has made it indispensable in both clinical practice and cardiovascular research (1, 2). Extraction of these parameters is typically performed on contiguous stacks of cine images acquired in short-axis orientation. For large-scale cohorts, such as those encountered in population imaging studies aimed at understanding the epidemiology of cardiovascular diseases, conventional – manual or semi-manual – segmentation methods for short-axis cine images are deemed impractical, primarily due to their labor-intensive nature and susceptibility to inter- and intra-observer variability. Moreover, suboptimal reproducibility across multi-center datasets may mask subtle variations in cardiac morphology and function across diverse populations. This is additionally complicated by variations in image quality, which may be adversely affected by diverse factors, including image artifacts associated with breathing or cardiac motion when ineffectively compensated for by electrocardiogram (ECG) gating. Overcoming these challenges demands advanced automated segmentation approaches and thorough quality control measures.

Automated segmentation techniques for CMR cine images have recently emerged through innovations in deep learning, notably the nnU-Net architecture (3, 4). These approaches now generate segmentation data with high robustness and achieve accuracies comparable to inter-observer variability on an average level, while helping to reduce biases (5, 6). Nonetheless, their use in large population studies still requires thorough quality control (7). When implemented effectively, their application in population imaging and epidemiology has enabled the creation of reliable CMR datasets that support the public health objective of disease prevention (8, 9).

The present study leverages data from two segmentation challenges (4, 5) and the prospective, multi-center NAKO to develop an image processing pipeline specifically tailored for short-axis cine CMR images. By integrating a deep learning model for automated segmentation with a comprehensive post-processing quality control framework it aims to generate a quality assured dataset of comprehensive CMR parameters that enables downstream analyses and ensures their integrity.

## Materials and Methods

### Ethics and participant consent

The NAKO Use and Access Committee approved this project based on the participants’ informed consent, its accordance with the aims of the NAKO, and ethical approval from the Ethics Committee of the Medical Faculty of Heidelberg University, Germany (S-972/2020, approved on February 3, 2021). This study conformed to the ethical guidelines of the 1964 Declaration of Helsinki and its later amendments.

### Project design

An overview of the design is given in **Figure 1**.

**Figure 1.**
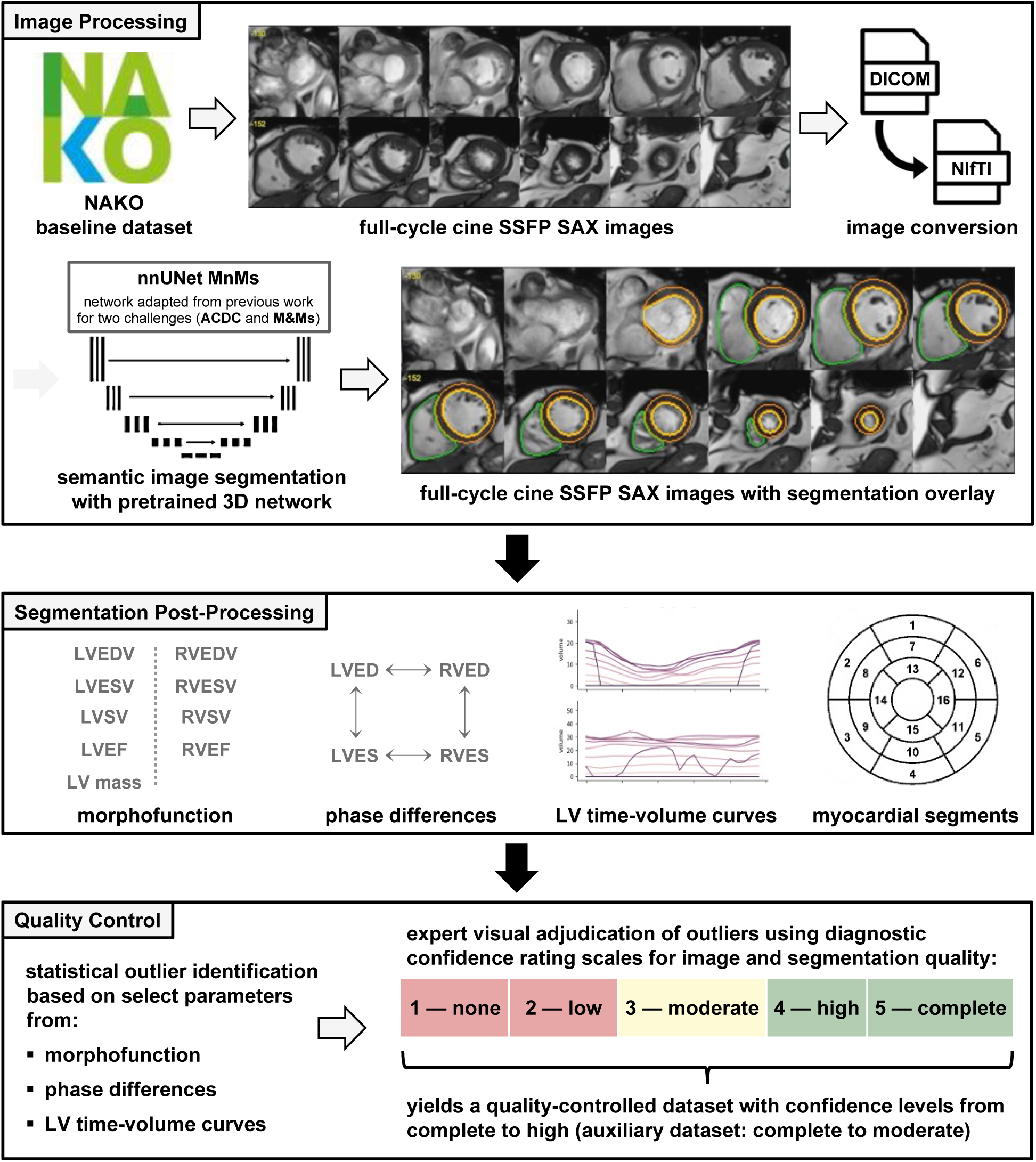
Overview of the study design, including the developed pipeline for image segmentation and quality control. *ACDC, Automated Cardiac Diagnosis Challenge; DICOM, Digital Imaging and Communications in Medicine; LV, left ventricle; LVED, left ventricular end diastole; LVEDV, left ventricular end diastolic volume; LVEF, left ventricular ejection fraction; LVES, left ventricular end systole; LVESV, left ventricular end systolic volume; LVSV, left ventricular stroke volume; M&Ms, Multi-Centre, Multi-Vendor and Multi-Disease Cardiac Segmentation Challenge; NIfTI, Neuroimaging Informatics Technology Initiative; RVED, right ventricular end diastole; RVEDV, right ventricular end diastolic volume; RVEF, right ventricular ejection fraction; RVES, right ventricular end systole; RVESV, right ventricular end systolic volume; RVSV, right ventricular stroke volume; SAX, short-axis; SSFP, steady-state free precession*

Our project was built on data from the NAKO, which is an ongoing, prospective, multicenter, population-based cohort study conducted by a network of 25 institutions at 18 regional examination sites in Germany. Its main objective is to investigate risk factors for the development of common chronic diseases such as cancer, diabetes, cardiovascular, neurodegenerative/psychiatric, respiratory, and infectious diseases (10). The baseline assessment was conducted between 2014 and 2019 and 205,415 participants from the general population aged 19 to 74 years were enrolled independently from known cardiovascular disease. The embedded NAKO MR imaging study, conducted at five dedicated imaging centers, enrolled 30,868 of these participants to receive a whole-body MR examination.

A detailed technical description of whole-body MR imaging in the NAKO has been published previously (11). MR imaging was performed on 3 T whole-body MR scanners (MAGNETOM Skyra, Siemens Healthineers, Erlangen, Germany) running an identical software version. The imaging program involved a cardiac assessment using functional and quantitative imaging techniques that included the acquisition of a steady-state free precession (SSFP) full-cycle cine image stack in the short-axis (SAX) orientation. By default, this consisted of 12 slices with 6 mm section thickness covering the heart from base to apex, with 25 phases evenly distributed over the cardiac cycle (11). All image acquisitions were performed following a standard operating procedure by radiologic technologists who were specifically trained and certified for the study. The cardiac planes were automatically planned using a vendor-provided software (Cardiac Dot Engine, Siemens Healthineers, Erlangen, Germany). The technologists were instructed to repeat any protocol if anatomic coverage did not meet the SOP, if severe image artifacts occurred, or if the image quality was unsatisfactory for other reasons (12).

For our project, we considered all available data at the time of investigation, which comprised baseline whole-body MR examinations from 30,868 participants (excluding data from participants who withdrew consent).

### Image processing

Short-axis cine images were available for 29,908 participants. They were converted from the DICOM (Digital Imaging and Communications in Medicine) format to the NIfTI (Neuroimaging Informatics Technology Initiative) format using an open source tool (13) and subsequently processed by a deep learning-based segmentation algorithm previously developed with the participation of authors of this paper (3, 4). It is built around two U-Net architectures; one that operates in two dimensions (to processes each slice separately) and another that operates in three dimensions (for additional exploitation of semantics across slices). Used in conjunction, the models are able to correct each other’s errors and perform better than either one performs individually. The algorithm output comprises full-cycle annotations for the endocardial and epicardial contours of the LV (together forming the myocardial segmentation) as well as the endocardial contour of the RV. As part of the published segmentation challenge, the models were trained on a public dataset that had been annotated by experts according to the guidelines by the Society for Cardiovascular Magnetic Resonance (14). A detailed technical description is provided in the original publication (3).

To determine the extent of differences across different annotation methods in terms of quantitative results, we initially applied the segmentation algorithm to a method development dataset of Cine SSFP SAX images from 11,050 NAKO participants. A random sample of 30 participants was drawn and processed in two software environments; syngo.via (Siemens Healthineers, Erlangen, Germany) and cvi42 (Circle Cardiovascular Imaging, Calgary, Canada), the contours were then manually corrected by two experienced annotators. A comparison of the target parameters showed that the variabilities between the two manual and algorithmic processing methods did not exceed that between the manual methods **(Supplemental Figure S1)**. Therefore, following finalization of the quality assurance methodology (detailed in the ‘Quality Assurance’ section below), we used the unmodified version of this validated algorithm for the current research project and applied it to the short-axis cine images in the baseline dataset. This process generated complete prediction data for 29,609 participants, while 299 participants (1.0 %) had incomplete data due to missing segmentations for one or more phases **(Figure 2)**.

**Figure 2.**
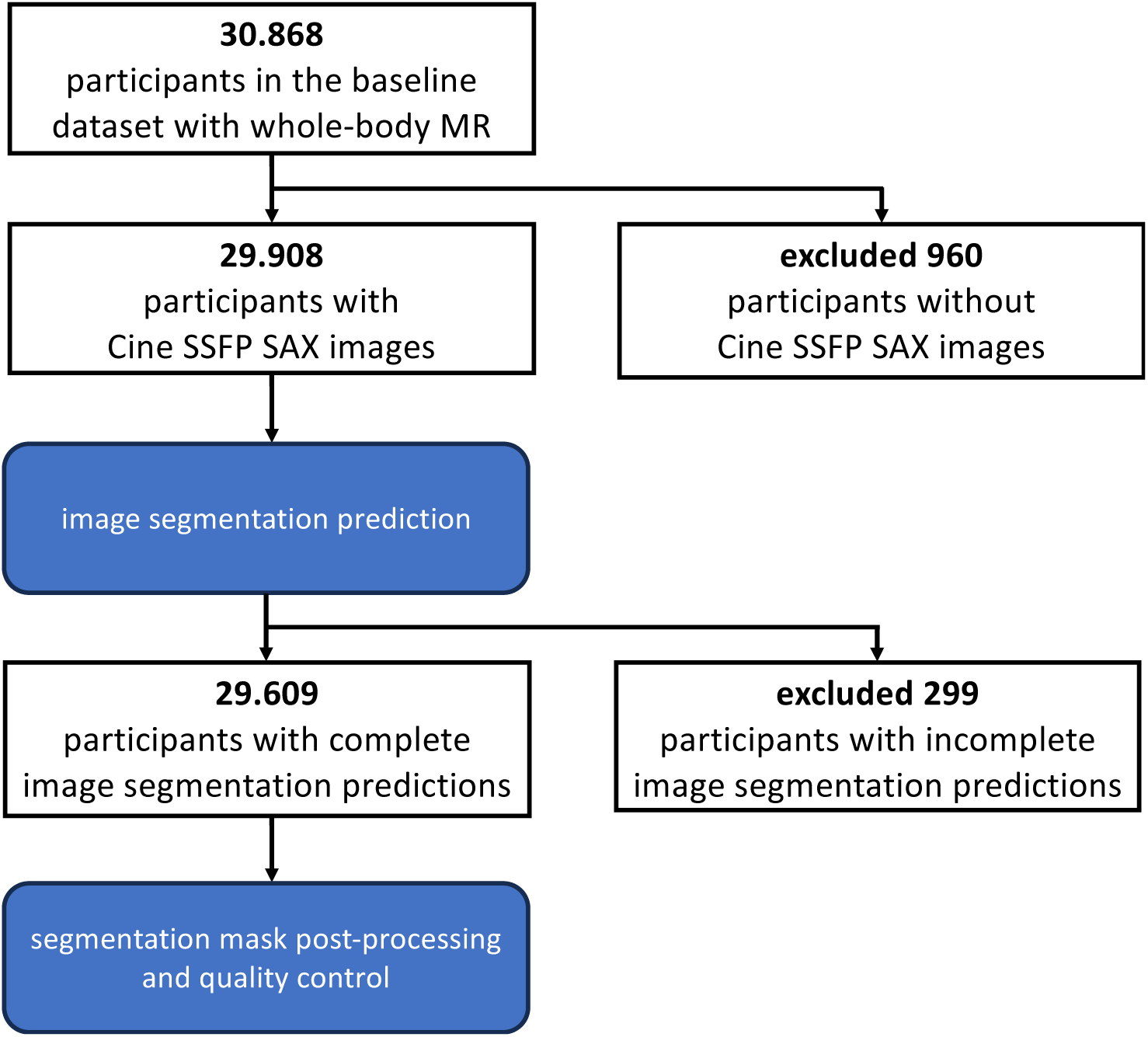
Flowchart of the study sample. *MR, magnetic resonance; SAX, short-axis; SSFP, steady-state free precession*

### Segmentation post-processing

The output of the segmentation algorithm was utilized to identify end diastolic and end systolic phases for both ventricles based on maximum and minimum volumes (t_LVED_, t_LVES_, t_RVED_ and t_RVES_). These cardiac phases were subsequently used to calculate inter-ventricular phase differences at end-diastole (dp_LVED/RVED_) and end-systole (dp_LVES/RVES_), as well as intra-ventricular phase differences between end-diastole and end-systole (dp_LVED/LVES_, dp_RVED/RVES_). Additionally, the following morphofunctional parameters were derived: ventricular end diastolic and end systolic volumes (LVEDV and LVESV, RVEDV and RVESV), ventricular stroke volumes (LVSV, RVSV), ventricular ejection fractions (LVEF, RVEF), left-ventricular myocardial mass (LV mass, calculated at end-diastole with a tissue density of 1.055 g/ml). Using a combination of temporal data and segmentation data, time-volume curves (TVC) for the LV endocardial blood pool were computed.

By further processing of the segmentation data, a 17-segment model for myocardial wall thickness at end-diastole was computed, following recommendations from the American Heart Association (AHA) (15). Further methodological details, including a schematic overview **(Supplemental Figure S2)**, are presented in the Supplemental Materials.

### Quality control

The results from the method development dataset were assessed to identify participants with outlying values: For the morphofunctional parameters LVEDV, LVESV, LV mass, RVEDV, RVESV—all with and without normalization to body surface area (BSA)—and LVEF and RVEF, the ten participants with the highest and lowest values were included in an exploratory sample for visual review. This sample comprised 139 distinct participants. A random selection of 200 participants was added and two expert readers (P.M.F, C.S.) jointly reviewed each of the 339 participants using an open-source image viewer with a dual-layout, showing the short-axis cine images with and without the segmentation overlay. Based on their findings related to cardiac structure representation and segmentation correctness, detailed criteria were established for two five-point rating scales of diagnostic confidence (5—complete confidence, 4—high confidence, 3—moderate confidence, 2—low confidence, 1—no confidence): A first rating scale for evaluating the image quality considering the presence and severity of image artifacts (especially from breathing or inconsistent ECG-synchronization), misalignment of the image stack from the intended short-axis orientation, shifts of slices along the short-axis, and missing slices. A second rating scale for evaluating the segmentation quality, focusing on the presence and severity of oversegmentation or undersegmentation errors, including transposition of segmentations into other organs or missing segmentations. Effects typically associated with the segmentation model—such as the potential “fraying” observed between segmentations of adjacent slices at the base or apex, attributable to interpolation or partial-volume effects—were classified as segmentation artifacts and exempted from penalization rather than being designated as segmentation errors. The score in the combined rating could only be as high as the initial image quality rating. Concurrently with this rating procedure, the criteria of the two rating scales were re-evaluated and refined to ensure appropriate rating assignments. The final rating scale is shown in **Supplemental Figure S3**, accompanied by additional explanatory notes.

The results from the entire baseline dataset were subsequently analyzed to identify outliers. For morphofunctional parameters (LVEDV, LVESV, LV mass, RVEDV, RVESV (all normalized to BSA), LVEF, and RVEF), complemented by the inter-ventricular difference in stroke volumes (dSV; calculated as ‘LVSV minus RVSV’), outlier values were defined as those exceeding or falling below 2.5 standard deviations from the gender-specific median. For inter-ventricular phase differences (dp_LVED/RVED_, dp_LVES/RVES_), outliers were defined as those exceeding two cardiac phases, whereas for intra-ventricular phase differences (dp_LVED/LVES_, dp_RVED/RVES_), they were defined as those shorter than six cardiac phases. For LV time-volume curves, outliers were identified as those with first mode scores exceeding the 91^st^ percentile. The identified participants were visually assessed by an expert reader (C.S.) using a local instance of the NORA imaging platform (16). A multi-layout was chosen showing the short-axis cine images with and without the segmentation overlay alongside long-axis images in two/three/four chamber views for reference **(Supplemental Figure S4)**. Comparing between multiple imaging planes supported differentiating between image artifacts and asynchronous cardiac motion patterns. There was no blinding to which parameters identified the participant as an outlier. From these final ratings, a quality-controlled dataset was constructed by excluding participants who received scores of ≤3 in either image quality or segmentation quality. For a more permissive alternative approach, an auxiliary dataset was constructed by excluding participants with scores of ≤2 in either rating.

### Statistics

Data on participants are presented as counts and percentages. Data on cardiac morphofunctional parameters are presented as continuous values. Fitted values for a generalized linear model with logit link were used to construct receiver operating characteristic (ROC) curves with corresponding areas under the curve (AUCs) for outcome exclusion for single outlier flags and their combinations. Differences in AUCs were compared by DeLong test. Differences in median values of morphofunctional parameters between included and excluded participants were compared using the Mann–Whitney U test. P-values <0.05 were considered to indicate statistical significance. Analyses used SAS version 9.4 (SAS Institute Inc., Cary, NC, USA) and R version 4.4.2 (R Foundation for Statistical Computing, Vienna, Austria).

## Results

### Outlier analysis: baseline dataset

Based on morphofunctional parameters, phases and time-volume curves, 5,082 (17.0 %) participants with complete segmentation data were identified as outliers. The majority of these were associated with a single outlier category, concerning 4,017 participants (79.0 % of all outliers), whereas 839 (16.5 %) and 226 (4.4 %) participants were associated with two and three outlier categories, respectively **(Figure 3)**. The visual assessment lead to inclusion rates from 35.1 % to 44.8 % per outlier category and from 8.6 % to 69.0 % per parameter **(Table 1)**. A total of 1,875 outliers were excluded based on image quality alone, followed by an additional 835 outliers excluded due to segmentation quality issues. Thus, 2,710 outliers (53.3 %) were excluded, resulting in 26,899 participants retained in the quality-controlled baseline dataset, which includes only cases with complete or high diagnostic confidence (inclusion rate: 90.8 %).

**Figure 3.**
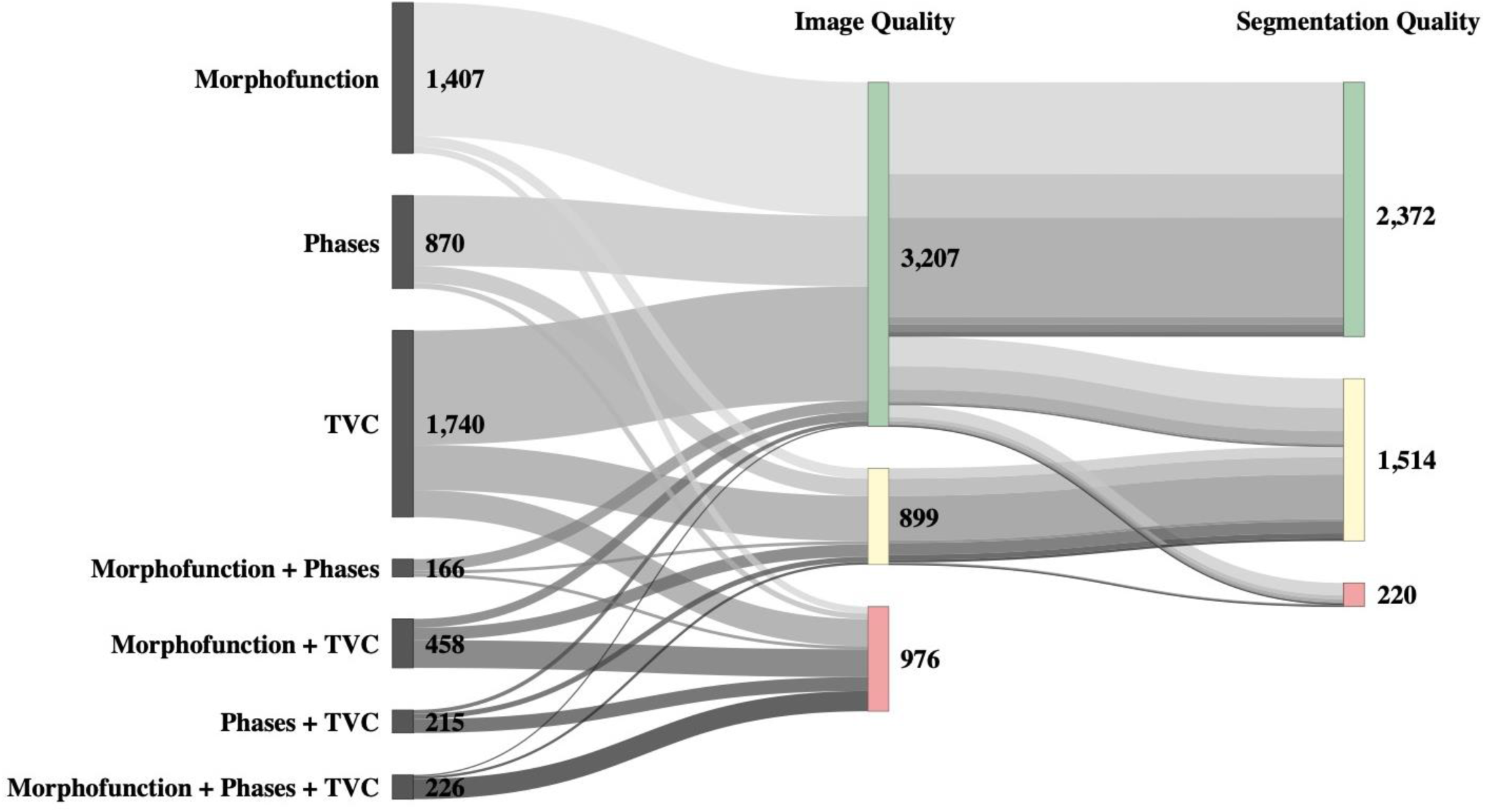
Sankey diagram illustrating multi-step quality control outcomes for CMR data from 5,082 participants. The first column groups participants into individual and combined outlier categories. The second and third columns display diagnostic confidence ratings for image and segmentation quality, respectively. Confidence levels are color-coded: green indicates complete or high confidence (ratings 5–4), blue indicates moderate confidence (3), and red indicates low or no confidence (2–1). *cardiac magnetic resonance (imaging); TVC, time-volume curves*

**Table 1.**
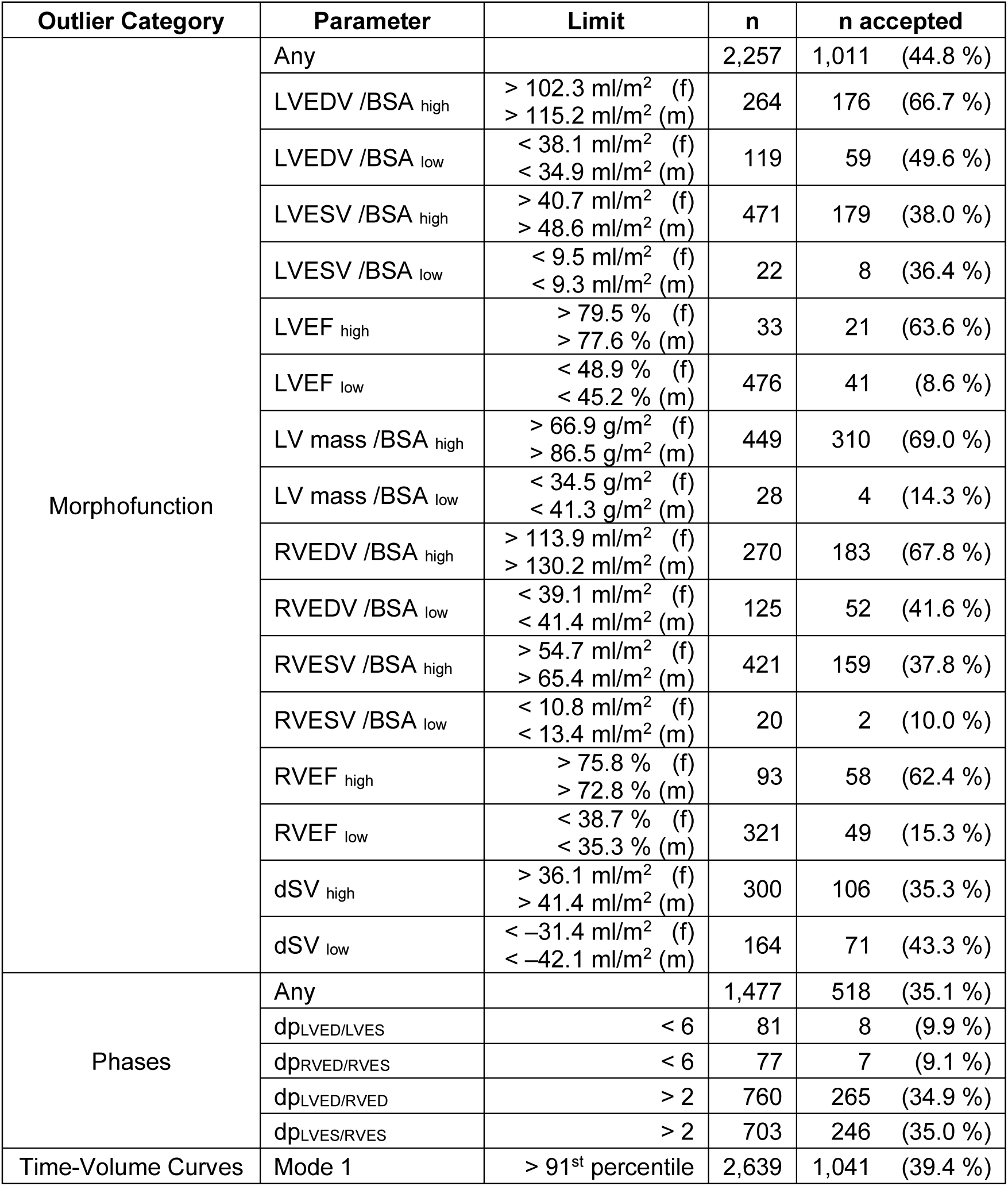
Results from the outlier analysis of the processed baseline dataset. For each of the parameters above*, n* specifies the number of participants identified as outliers, based on threshold limits normalized separately for females (f) and males (m), where applicable. *n accepted* specifies the number of participants with complete or high diagnostic confidence regarding data quality (ratings 5–4), along with the corresponding percentage relative to *n*. *BSA, body surface area; dp, difference in cardiac phase; dSV, difference in ventricular stroke volumes; LV, left ventricle; LVEDV, left ventricular end diastolic volume; LVEF, left ventricular ejection fraction; LVESV, left ventricular end systolic volume; RVEDV, right ventricular end diastolic volume; RVEF, right ventricular ejection fraction; RVESV, right ventricular end systolic volume*

| Outlier Category | Parameter | Limit | n | n accepted |
| --- | --- | --- | --- | --- |
| Morphofunction | Any |  | 2,257 | 1,011 (44.8 %) |
|  | LVEDV /BSA<sub>high</sub> | > 102.3 ml/m<sup>2</sup> (f)<br>> 115.2 ml/m<sup>2</sup> (m) | 264 | 176 (66.7 %) |
|  | LVEDV /BSA<sub>low</sub> | < 38.1 ml/m<sup>2</sup> (f)<br>< 34.9 ml/m<sup>2</sup> (m) | 119 | 59 (49.6 %) |
|  | LVESV /BSA<sub>high</sub> | > 40.7 ml/m<sup>2</sup> (f)<br>> 48.6 ml/m<sup>2</sup> (m) | 471 | 179 (38.0 %) |
|  | LVESV /BSA<sub>low</sub> | < 9.5 ml/m<sup>2</sup> (f)<br>< 9.3 ml/m<sup>2</sup> (m) | 22 | 8 (36.4 %) |
|  | LVEF<sub>high</sub> | > 79.5 % (f)<br>> 77.6 % (m) | 33 | 21 (63.6 %) |
|  | LVEF<sub>low</sub> | < 48.9 % (f)<br>< 45.2 % (m) | 476 | 41 (8.6 %) |
|  | LV mass /BSA<sub>high</sub> | > 66.9 g/m<sup>2</sup> (f)<br>> 86.5 g/m<sup>2</sup> (m) | 449 | 310 (69.0 %) |
|  | LV mass /BSA<sub>low</sub> | < 34.5 g/m<sup>2</sup> (f)<br>< 41.3 g/m<sup>2</sup> (m) | 28 | 4 (14.3 %) |
|  | RVEDV /BSA<sub>high</sub> | > 113.9 ml/m<sup>2</sup> (f)<br>> 130.2 ml/m<sup>2</sup> (m) | 270 | 183 (67.8 %) |
|  | RVEDV /BSA<sub>low</sub> | < 39.1 ml/m<sup>2</sup> (f)<br>< 41.4 ml/m<sup>2</sup> (m) | 125 | 52 (41.6 %) |
|  | RVESV /BSA<sub>high</sub> | > 54.7 ml/m<sup>2</sup> (f)<br>> 65.4 ml/m<sup>2</sup> (m) | 421 | 159 (37.8 %) |
|  | RVESV /BSA<sub>low</sub> | < 10.8 ml/m<sup>2</sup> (f)<br>< 13.4 ml/m<sup>2</sup> (m) | 20 | 2 (10.0 %) |
|  | RVEF<sub>high</sub> | > 75.8 % (f)<br>> 72.8 % (m) | 93 | 58 (62.4 %) |
|  | RVEF<sub>low</sub> | < 38.7 % (f)<br>< 35.3 % (m) | 321 | 49 (15.3 %) |
|  | dSV<sub>high</sub> | > 36.1 ml/m<sup>2</sup> (f)<br>> 41.4 ml/m<sup>2</sup> (m) | 300 | 106 (35.3 %) |
|  | dSV<sub>low</sub> | < -31.4 ml/m<sup>2</sup> (f)<br>< -42.1 ml/m<sup>2</sup> (m) | 164 | 71 (43.3 %) |
| Phases | Any |  | 1,477 | 518 (35.1 %) |
|  | dp<sub>LVED</sub>/LVES | < 6 | 81 | 8 (9.9 %) |
|  | dp<sub>RVED</sub>/RVES | < 6 | 77 | 7 (9.1 %) |
|  | dp<sub>LVED</sub>/RVED | > 2 | 760 | 265 (34.9 %) |
|  | dp<sub>LVES</sub>/RVES | > 2 | 703 | 246 (35.0 %) |
| Time-Volume Curves | Mode 1 | > 91<sup>st</sup> percentile | 2,639 | 1,041 (39.4 %) |

Under the more permissive alternative approach, where participants with ratings of ‘3—moderate confidence’ were retained as well, these numbers changed notably: Then, 976 outliers were excluded due to image quality and 220 due to segmentation issues, totaling 1,196 exclusions (23.5 %). This resulted in an auxiliary baseline dataset of 28,413 participants (inclusion rate: 96.0 %).

Among the single outlier categories, TVC showed the best discriminative ability to distinguish exclusion from inclusion compared to morphofunction criteria (difference in AUC: 0.166, 95%CI: [0.149, 0.182], p<0.001) or phase difference criteria (difference in AUC: 0.140 [0.119, 0.160], p<0.001). Among the combination of two categories, the combination of TVC and phase difference criteria showed the best discriminative ability, compared to the combination of morphofunction criteria and TVC (difference in AUC: 0.019 [0.010, 0.028], p<0.001) or the combination of morphofunction and phase difference criteria (difference in AUC: 0.162 [0.145, 0.179], p<0.001). Moreover, the combination of TVC and phase difference criteria showed better discriminative ability than TVC alone (difference in AUC: 0.044 [0.039, 0.050], p<0.001). The combination of all three categories showed best discriminative ability overall (difference in AUC to the combination of TVC and phase differences: 0.020 [0.014, 0.026], p<0.001) **(Figure 4)**.

**Figure 4.**
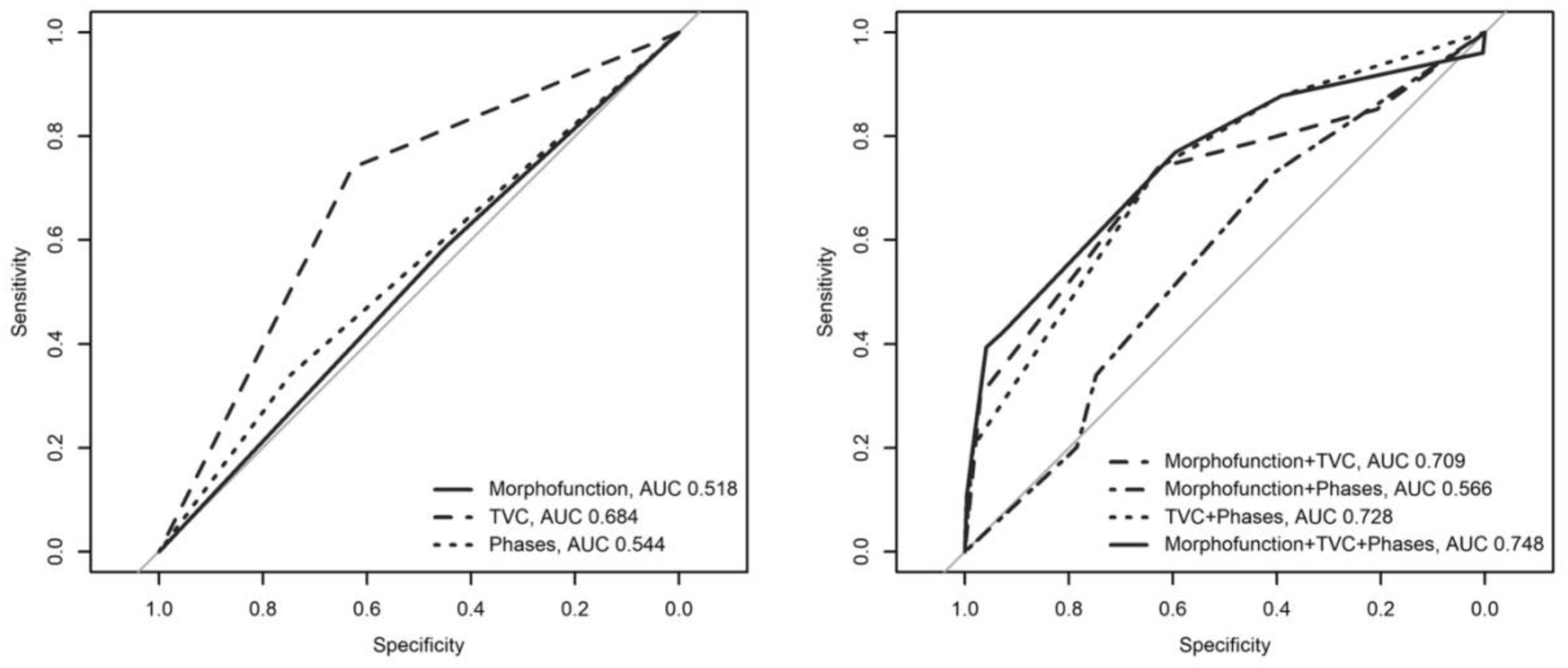
Receiver operating characteristics curves (ROC) and areas under the curve (AUCs) for outcome exclusion for single outlier categories and their combinations. *TVC, time-volume curves*

### Comparison of ventricular structure and function between included and excluded participants

The distribution patterns of morphofunctional parameters for excluded and included outliers were similar in shape **(Figure 5)**. Although statistically significant, the differences in their median values were generally small: In excluded participants, decreases were seen for end diastolic volumes (LVEDV: – 1.5 ml/m^2^, RVEDV: – 1.9 ml/m^2^, both p < 0.001), stroke volumes (LVSV: – 3.6 ml/m^2^, RVSV: – 4.3 ml/m^2^, both p < 0.001), ejection fractions (LVEF: – 2.3 %, RVEF: – 4.3 %, both p < 0.001), and LV mass (– 3.5 g/m^2^, p < 0.001), whereas increases were seen in end systolic volumes (LVESV: + 1.0 ml/m^2^, RVESV: + 2.0 ml/m^2^, both p < 0.001).

**Figure 5.**
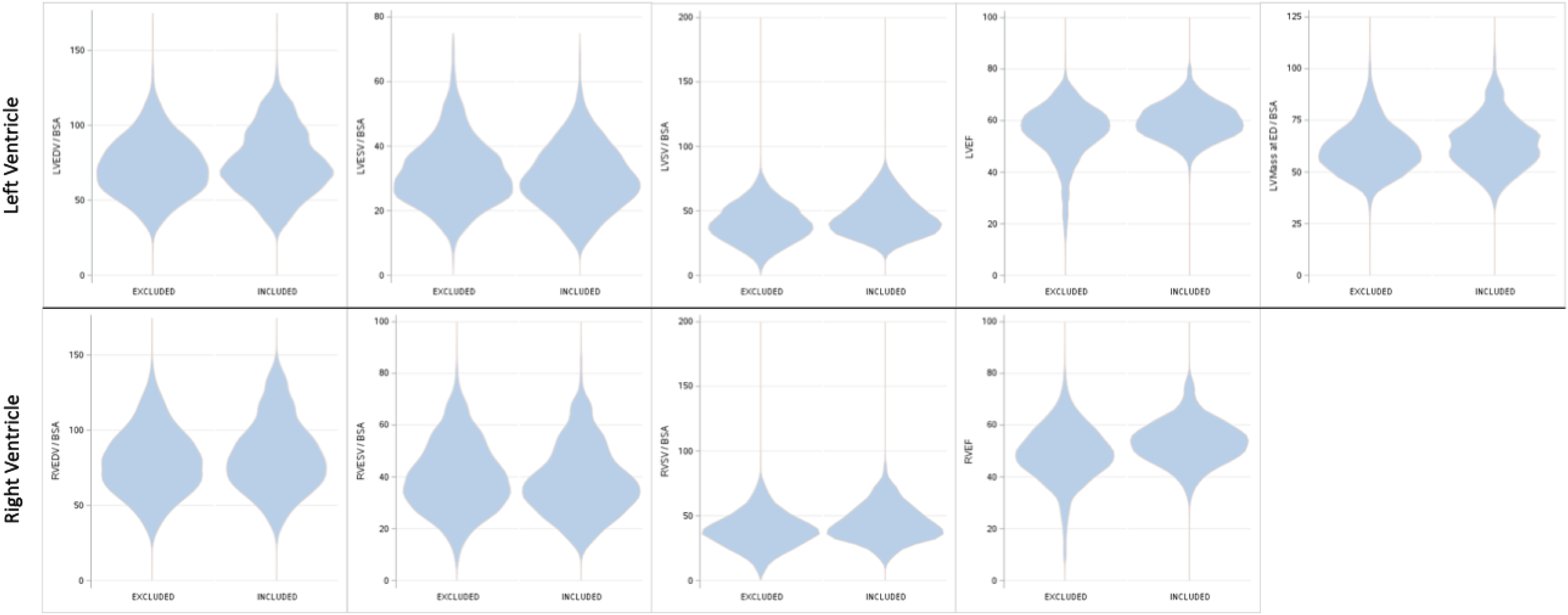
Violin plots illustrating the distribution of morphofunctional parameters between included and excluded participants. *BSA, body surface area; ED, end diastole; LV, left ventricle; LVEDV, left ventricular end diastolic volume; LVEF, left ventricular ejection fraction; LVESV, left ventricular end systolic volume; LVSV, left ventricular stroke volume; RVEDV, right ventricular end diastolic volume; RVEF, right ventricular ejection fraction; RVESV, right ventricular end systolic volume; RVSV, right ventricular stroke volume*

## Discussion

This study on cardiac MR imaging data from the baseline sample of the NAKO Health Study introduced a pipeline for the automated segmentation of short-axis cine images together with a post-processing quality control workflow. The process enabled the extraction of quantitative parameters and the exclusion of participants with suboptimal image or segmentation quality. Consequently, we compiled a comprehensive dataset of cardiac structure and function from a large population-based cohort (n = 26,899 with complete or high diagnostic confidence and n = 28,413 with complete to moderate diagnostic confidence), providing a robust foundation for further analyses.

A challenge in segmenting large image datasets and performing quality control is the complex nature of potential interfering factors. Image quality may be degraded by a diverse range of imaging artifacts, which may arise from the imaging equipment and its operation, from the examined person and their conscious or unconscious actions, or from other possibly unidentified factors impeding the imaging process. Segmentation quality may likewise be compromised by many causes, including suboptimal input data or intrinsic problems within the segmentation model. Identifying all relevant confounders requires considerable effort and may not be feasible, yet identifying and filtering out acquisitions with low quality is paramount to ensure the integrity of downstream analyses. In our study, we approached this task by processing the unfiltered image data in the form of short-axis cine images with a segmentation model on the nnU-Net architecture that was previously validated to yield accurate predictions with a high robustness (5, 6). Following the rationale that significantly impaired images would influence segmentation quality or, if accurately segmented, exhibit implausible parameters of cardiac morphofunction, we appended the quality control process after the segmentation, as opposed to pre-filtering participants with subpar short-axis cine images. This allowed us to perform an outlier analysis on the segmentation results and retrospectively identify participants with either impaired image quality, segmentation quality, or both. Our results underline the importance of expert visual quality controls alongside statistical methods in cardiac imaging, as exclusions could not be reliably identified through statistical criteria alone. This was further illustrated by the comparable parameter distribution shapes between included and excluded participants. Although the exclusion rate of 9.2 % in the main quality-controlled dataset may appear relatively high, it reflects the application of strict quality standards—arguably exceeding those typically employed in clinical practice. For analyses requiring less stringent quality control, the auxiliary dataset, which has a lower exclusion rate of 4.0 %, may offer a feasible alternative.

Other studies have explored deeper preprocessing image quality control methods. In the UK Biobank (UKBB), these included techniques based on intricate knowledge of MR physics for detecting issues like asynchronicity between slices or mistriggering due to inadequate ECG gating (17–19). Related methods could sometimes be utilized to correct cine images from the UKBB dataset using techniques like k-space reconstruction (20, 21). Adapting and validating such algorithms for new datasets requires substantial effort. If only single slices are affected in SAX stacks, a simpler alternative approach could involve omitting the affected slices followed by data interpolation from adjacent slices, thereby limiting the introduced error and enabling re-inclusion into the quality-controlled dataset. Preprocessing image quality control based on image sharpness, signal-to-noise ratio, and other general quantitative metrics showed only moderate performance when applied to cardiac cine images from the NAKO (12, 22).

Our study focused on morphofunctional analysis, particularly in the context of end-diastolic (ED) and end-systolic (ES) phases, rather than on shape-model accuracy. Consequently, our methodology was not optimized to produce anatomically shaped segmentations, and interpolation artifacts were accepted as long as they fell within the space of plausible annotations. While these artifacts may appear unfamiliar to the human eye, they do not inherently compromise the primary morphofunctional endpoints of this study, as different analytical goals impose different requirements on segmentation accuracy. This is particularly relevant for the basal slices, where the distinction between atrial and ventricular borders is inherently challenging in short-axis images, and complicates distinguishing between segmentation inaccuracies and true algorithmic failures. While these inaccuracies could potentially average out at the whole-sample level, caution is warranted when interpreting individual participant data, even when segmentations receive high-quality rating. The benefit of retraining the segmentation model in this regard will likely be limited without incorporating long-axis cine images for improved delineation of the atrioventricular borders.

Our study has additional limitations. Notably, we did not assess segmentation quality independently of image quality. This decision was based on several factors. First, evaluating segmentation quality in isolation is inherently challenging, as it is closely tied to the underlying image. For instance, minor motion artifacts from ECG mistriggering or breathing could lead to an apparent segmentation deficiency. Additionally, given our primary objective of excluding insufficient data from the final sample, segmentation quality in isolation was not a determining factor. Regardless of the cause, our quality control process aimed to exclude participants with deficiencies in image quality or segmentation quality, supporting the creation of a reliable dataset. Even with the current system, participants suitable for model retraining can still be identified, specifically those with adequate image quality scores but insufficient segmentation quality scores. Furthermore, we did not perform a connected component analysis on the predicted segmentations. While this is likely not a problematic factor for morphofunctional parameters alone, it would need to be conducted before shape model analyses on the segmentation data could be reliably performed. Lastly, the validity of the derived LV parameters is expected to be higher than that of RV parameters, as MR-based RV measurements are considered to be more accurate when performed on cine images in the body axial rather than the cardiac short-axis orientation (23, 24).

To conclude, the presented pipeline enabled automated segmentation of short-axis cine cardiac MR images from the baseline cohort of the NAKO Health Study. By incorporating a comprehensive quality control process, it generated a comprehensive dataset of quantitative information on cardiac structure and function that is suitable for downstream analyses.

## Acknowledgments

This project was conducted with data (application no. NAKO-249, NAKO-744) from the German National Cohort (NAKO) (www.nako.de). The NAKO is funded by the Federal Ministry of Education and Research (BMBF) [project funding reference numbers: 01ER1301A/B/C, 01ER1511D, 01ER1801A/B/C/D, and 01ER2301A/B/C], the federal states of Germany, the Helmholtz Association, the participating universities, and the institutes of the Leibniz Association. We thank all participants who took part in the NAKO and the staff in this research initiative.

P.M.F. was funded by the Kaltenbach Scholarship of the German Heart Foundation.

## Data availability

Access to and use of NAKO data and biosamples can be obtained via an electronic application portal (https://transfer.nako.de). This includes the data generated in the current study.

## Abbreviations

AHA: American Heart Association
AUC: area under the curve
BSA: body surface area
CMR: cardiac magnetic resonance (imaging)
DICOM: Digital Imaging and Communications in Medicine
dp: difference in cardiac phase
dSV: difference in ventricular stroke volumes (‘LVSV minus RVSV’)
FHS: Framingham Heart Study
LV: left ventricle
LVEDV: left ventricular end diastolic volume
LVEF: left ventricular ejection fraction
LVESV: left ventricular end systolic volume
LVSV: left ventricular stroke volume
MESA: Multi-Ethnic Study of Arteriosclerosis
MR: magnetic resonance (imaging)
NAKO: German National Cohort
NIfTI: Neuroimaging Informatics Technology Initiative
ROC: receiver operating characteristic
RV: right ventricle
RVEDV: right ventricular end diastolic volume
RVEF: right ventricular ejection fraction
RVESV: right ventricular end systolic volume
RVSV: right ventricular stroke volume
SAX: short-axis
SHIP: Study of Health in Pomerania
SSFP: steady-state free precession
TVC: time-volume curves
UKBB: UK Biobank

