## Supplemental Materials for "Cardiac Magnetic Resonance Imaging in the German National Cohort: Automated Segmentation of Short-Axis Cine Images and Post-Processing Quality Control"

**Supplemental Figure S1.** Comparison of measurements obtained from expert segmentations using two systems (syngo.via and cvi42) and algorithmic processing in a random sample of 30 participants. The results show that **(a)** differences, illustrated by Bland-Altman plots, and **(b)** correlations, shown in scatter plots, between manual and automated measurements fall within the range of inter-reader variability observed in manual assessments.

*LV, left ventricle; LVEDV, left ventricular end diastolic volume; LVESV, left ventricular end systolic volume; RVEDV, right ventricular end diastolic volume; RVESV, right ventricular end systolic volume*

**Supplemental Figure S1.** (continued)

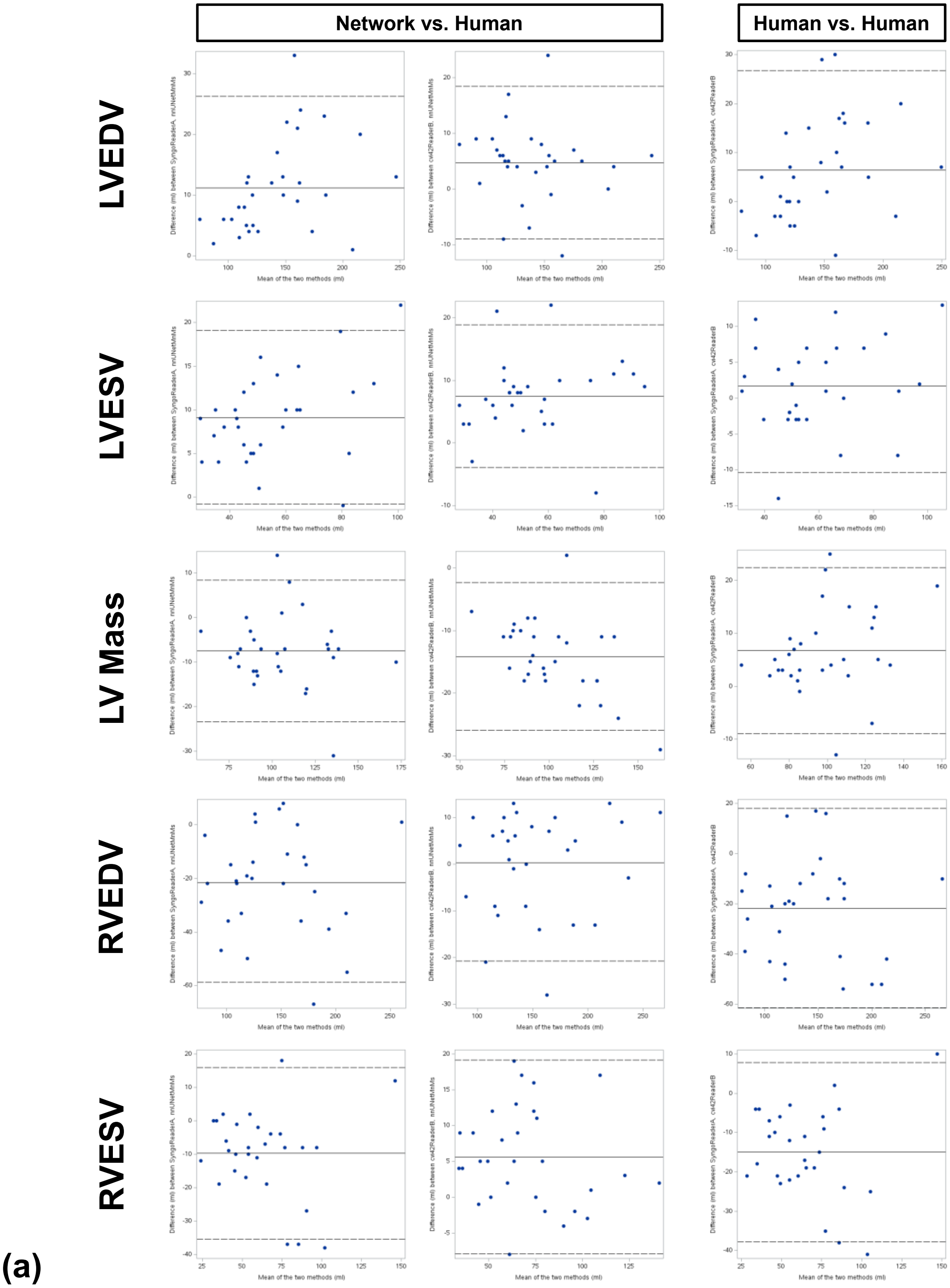

**Supplemental Figure S1.** (continued)

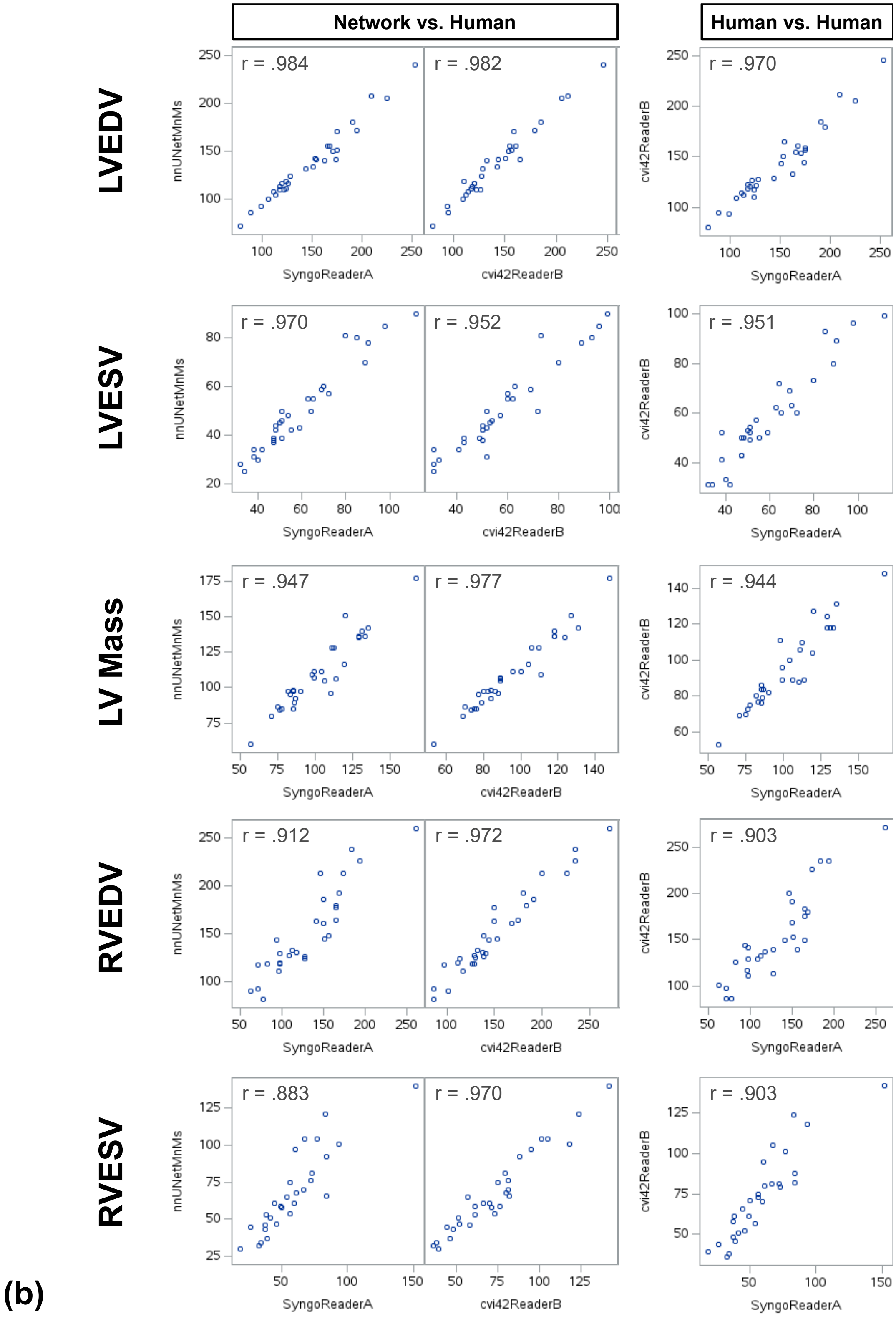

**Computation of Myocardial Wall Thickness**

A 17-segment AHA model for myocardial wall thickness at end-diastole was derived from the segmentation data. A schematic overview of the computation is shown in **Supplemental Figure S2**, with individual steps detailed below:

1. The volume of the LV mask was computed for all time steps and the time step with the largest volume was selected for further analysis.
2. The short-axis slices in the selected volume that contained any LV mask and any myocardial mask were divided equally into apical, mid-cavity, and basal thirds. This division could result in fractional assignments. For example, with ten slices, slices 1-3 are classified as 100 % apical, slice 4 as 33 % apical and 67 % mid-cavity, slices 5-6 as 100 % mid-cavity, slice 7 as 67 % mid-cavity and 33 % basal, and slices 8-10 as 100 % basal, such that each region (apical, mid-cavity, and basal) encompasses exactly 3⅓ slices.
3. For each such slice, endocardial and epicardial contour points, collectively forming the myocardial contour points, were identified: First, the contours of the myocardial mask were computed using the 'marching squares' methods as implemented in scikit-image[1]. Second, the center of the myocardium was estimated as the average of the myocardial point coordinates. Third, points were classified as inner or outer based on angle criteria: If the angle between the normal of the contour at a given point and the vector from the center to

[1] van der Walt S, Schönberger JL, Nunez-Iglesias J, Boulogne F, Warner JD, Yager N, et al. scikit-image: image processing in Python. PeerJ. 2014 2014/06/19;**2**:e453.

that point exceeded 90°, the point was considered an inner point, otherwise an outer point.

4. Myocardial thickness per outer point was calculated as the distance to the closest inner point.
5. The outer myocardial points were assigned to the corresponding myocardial segments. For that, the angles of all the vectors from the aforementioned myocardial center to the outer points were computed as well as the touching points between the myocardium and the RV (to determine the septum). The outer myocardial points belonging to the septum were further subdivided into the corresponding two septal segments in the basal and mid-cavity slices. The remaining outer points were subdivided into three (apical) or five (basal, mid-cavity) segments as per the AHA definition[2]. All subdivisions were based on the aforementioned angles. Slices in which more than 50% of the myocardial circumference was absent at end diastole—typically occurring at the left ventricular outflow tract—were excluded, whereas slices retaining more than 50% of the circumferential coverage were retained. However, to avoid bias in wall thickness estimation, 4.5 mm on either side of the “gaps” (i.e., the outflow tract) were removed.

---

[2] Cerqueira MD, Weissman NJ, Dilsizian V, Jacobs AK, Kaul S, Laskey WK, et al. Standardized myocardial segmentation and nomenclature for tomographic imaging of the heart. A statement for healthcare professionals from the Cardiac Imaging Committee of the Council on Clinical Cardiology of the American Heart Association. Circulation. 2002 Jan 29;**105**(4):539-42.

6. The segment thickness was then computed as the average of the myocardial thicknesses in that segment, also taking into account the fractional assignments of slices to apical, mid-cavity and basal as explained in step 2.

**Supplemental Figure S2.** Schematic overview of the myocardial wall thickness computation.

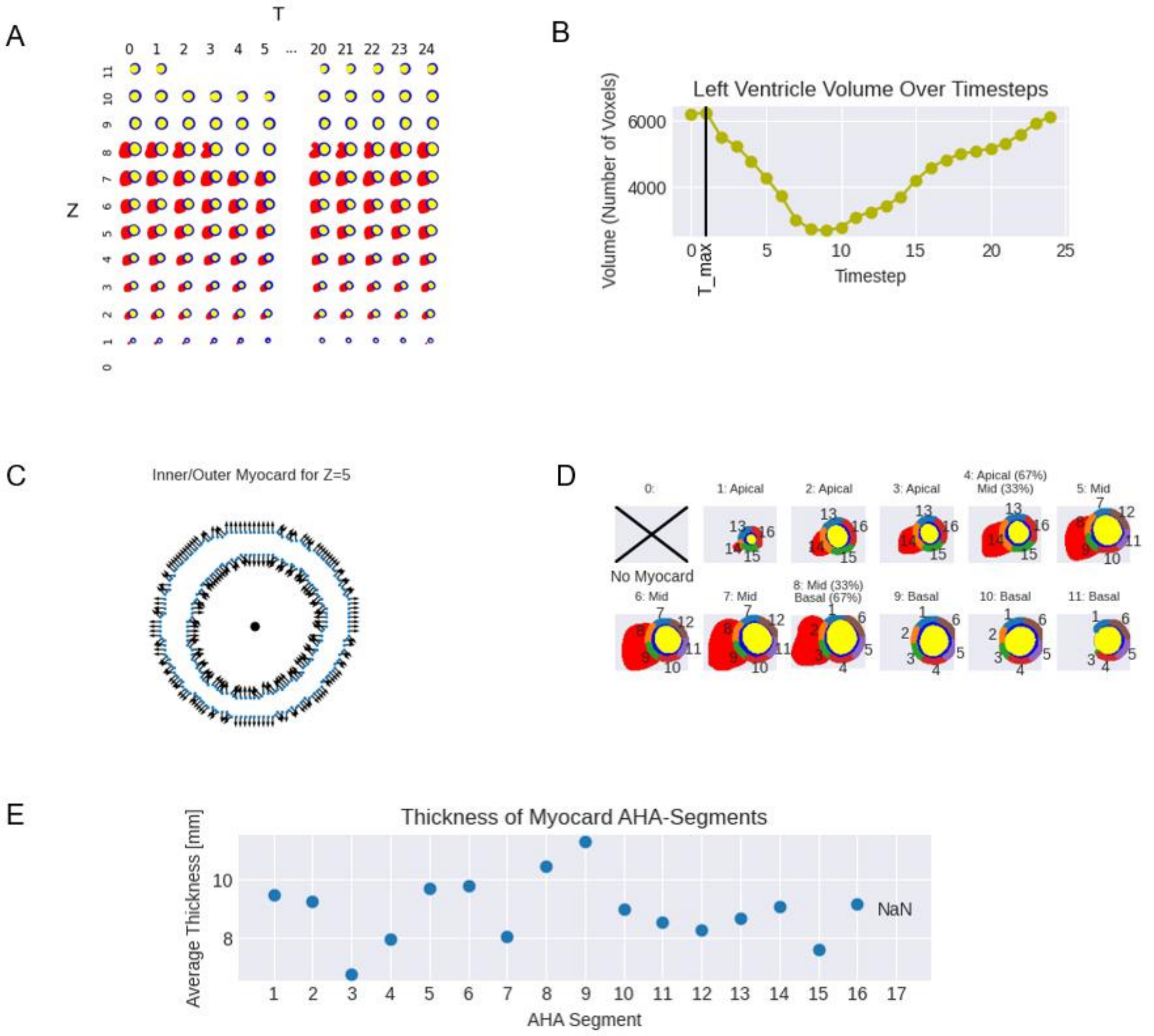


**A:** Segmentations across all phases and slices as input. **B:** Selection of the phase with maximum left ventricular volume. **C:** Identification of endocardial and epicardial contour points. **D:** Assignment of points to AHA segments. **E:** Computation of myocardial thickness per AHA segment as final output.

**Supplemental Figure S3.** The rating scales employed for the visual evaluation of the cardiac short-axis cine images during the quality control reads

| | 1—no confidence<br>very limited or no usable information on cardiac morphofunction | 2—low confidence<br>limited information on cardiac morphofunction | 3—moderate confidence | 4—high confidence<br>minor flaws unlikely to have a sizable influence the endpoints* | 5—complete confidence<br>no relevant flaws in relation to the endpoints* |
|---|---|---|---|---|---|
| Image Quality | Would not use clinically other than for crude visual estimations, if at all possible. | | Would use clinically with caution. | Would use clinically. | |
| | - Major image artifacts affecting the ventricles (flow artifacts, pulsation artifacts; relevant structures in larger parts indiscernible): ≥4 affected z-slices on a ventricular level<br>- Major motion artifacts (trigger artifacts/ asynchronous slices/ breathing artifacts; end-systole and end-diastole mostly indiscernible/not depicted): ≥4 affected z-slices on a ventricular level unassessable<br>- Imaging plane completely incorrect (e.g., LAX or random)<br>- ≥3 z-slices relevantly shifted (or swapped) along the z-axis<br><br>—> **≥4 z-slices are not assessable.** | - Medium image artifacts affecting the ventricles (flow artifacts, pulsation artifacts; relevant structures in relevantly (medium) sized areas or portions indiscernible): ≥4 affected z-slices on a ventricular level<br>OR<br>Major image artifacts affecting the ventricles (flow artifacts, pulsation artifacts; relevant structures in larger parts indiscernible): 2-3 affected z-slices on a ventricular level<br>- Medium motion artifacts (trigger artifacts/ asynchronous slices/ breathing artifacts; end-systole and end-diastole partly indiscernible/not strictly depicted/partly off): ≥4 affected z-slices on a ventricular level<br>OR<br>Major motion artifacts (trigger artifacts/ asynchronous slices/ breathing artifacts; end-systole and end-diastole mostly indiscernible/off): 2-3 affected z-slices on a ventricular level<br>- Imaging plane suboptimal (majorly misaligned with SAX)<br>- 2 z-slices relevantly shifted (or swapped) along the z-axis<br>- Missing z-slice at the basal or apical end: ≥2 (generally due to abort)<br><br>—> **Up to 3 z-slices are not assessable.** | - Medium image artifacts affecting the ventricles (flow artifacts, pulsation artifacts; relevant structures in relevantly (medium) sized areas or portions indiscernible): ≤3 affected z-slices on a ventricular level<br>OR<br>Major image artifacts affecting the ventricles (flow artifacts, pulsation artifacts; relevant structures in larger parts indiscernible): ≤1 affected z-slices on a ventricular level<br>- Medium motion artifacts (trigger artifacts/ asynchronous slices/ breathing artifacts; end-systole and end-diastole party indiscernible/not strictly depicted/partly off): ≤3 affected z-slices on a ventricular level<br>OR<br>Major motion artifacts (trigger artifacts/ asynchronous slices (including incomplete contraction)/ breathing artifacts; end-systole and end-diastole mostly indiscernible/off): ≤1 affected z-slices on a ventricular level<br>- 1 z-slice relevantly shifted (or swapped) along the z-axis<br>- Imaging plane suboptimal (medium misaligned with SAX)<br>- Missing z-slice at the basal or apical end: ≤1<br><br>—> **Up to 1 z-slice is not assessable.** | - Minor image artifacts affecting the ventricles (flow artifacts, pulsation artifacts; relevant structures reasonably discernible at ES/ED): any number of z-slices<br>- Minor motion artifacts (trigger artifacts/ asynchronous slices (including incomplete contraction)/ breathing artifacts; relevant structures reasonably discernible at ES/ED): any number of z-slices<br>- Shifting along the x/y axis within the imaging plane (up/down, left/right): any number of slices (acceptable for morphofunction, although not for shape analysis)<br>- Imaging plane suboptimal (minorly misaligned with SAX) | - No or very subtle image artifacts relevantly interfering with the depiction of the ventricles (image artifacts outside the ventricles may be present; e.g., pulmonary, abdominal)<br>- No or very subtle motion artifacts (trigger artifacts/ asynchronous slices/ breathing artifacts) interfering with the depiction of the contraction cycle<br>- Full depiction of the ventricles (no missing slices) |
| | - Incomplete capture of the cardiac cycle (fewer than 25 timepoints; less than half) [although strictly could include all relevant timepoints; affects only cases with complete processing failure; often 17 timepoints.] | - Incomplete capture of the cardiac cycle (fewer than 25 timepoints; but more than half) [although strictly could include all relevant timepoints; affects only cases with complete processing failure; often 17 timepoints.] | - More than one full cardiac cycle captured (technically incorrect image capture but usable under special consideration; however, not in this study). | | |
| | **— Stop —** | | **↙ Continue to Segmentation Quality ↘**<br>**(cannot be better than image quality rating)** | | |

**Supplemental Figure S3.** (continued)

| | 1—no confidence<br>very limited or no usable information on cardiac morphofunction | 2—low confidence<br>limited information on cardiac morphofunction | 3—moderate confidence | 4—high confidence<br>minor flaws unlikely to have a sizable influence the endpoints* | 5—complete confidence<br>no relevant flaws in relation to the endpoints* |
|---|---|---|---|---|---|
| | Would not use clinically other than for crude visual estimations, if at all possible. | | Would use clinically with caution. | Would use clinically. | |
| **Segmentation Quality** | - Segmentation failure: Any transposition into a non-adjacent structure (e.g., stomach), independent of timepoint/volumes<br>- Segmentation failure: Any contour missing completely in all z-slices | - Segmentation failure: Major oversegmentation into a closely adjacent structure (e.g., neighboring cardiac chamber, epicardial fat, pericardial fat) outside major image artifacts with obviously large effect on relevant* volumes<br>- Segmentation failure: Major undersegmentation outside major image artifacts with obviously large effect on relevant* volumes | - Segmentation failure: Medium oversegmentation into a closely adjacent structure (e.g., neighboring cardiac chamber, epicardial fat, pericardial fat) outside major image artifacts with potentially non-negligible effect on relevant* volumes (per subjective assessment)<br>- Segmentation failure: Medium undersegmentation (e.g., incomplete apical LV ring despite thick myocardium) outside major image artifacts with potentially non-negligible effect on relevant* volumes (per subjective assessment) | - Segmentation failure: Minor oversegmentation into a closely adjacent structure (e.g., neighboring cardiac chamber, epicardial fat, pericardial fat) outside major image artifacts with subjectively negligible/miniscule or very small effect on relevant* volumes<br>- Segmentation failure: Minor undersegmentation (e.g., incomplete apical LV ring despite thick myocardium) outside major image artifacts with subjectively negligible/miniscule or very small effect on relevant*volumes<br>- Segmentation training effect: 'Reversed Basal RA Build-Up or Build-Down' (expected in ~50% of cases) as long as subjectively minimal impact on relevant RV volumes | - Expert-like segmentation quality for all three contours.<br>- Segmentation artifact: Frayed basal RV contour (RV/RA border) due to interpolations<br>- Segmentation artifact: Incomplete basal or apical LV ring due to interpolations and/or partial-volume effects of myocardium and/or thin myocardium |

*The image quality rating was performed first and especially considered included motion artifacts from breathing or inconsistent ECG-synchronization, misalignment from the intended short-axis orientation, and missing slices. The segmentation quality rating combined image and segmentation quality, addressing errors such as oversegmentation or undersegmentation, while excluding artifacts inherently associated with the segmentation method and its handling of interpolation or partial-volume effects. The combined rating could only be as high as the initial image quality rating.*

**Supplemental Figure S4.** The image displays the NORA image viewer and the layout selected for visually evaluating cardiac short-axis cine images during the quality control reads

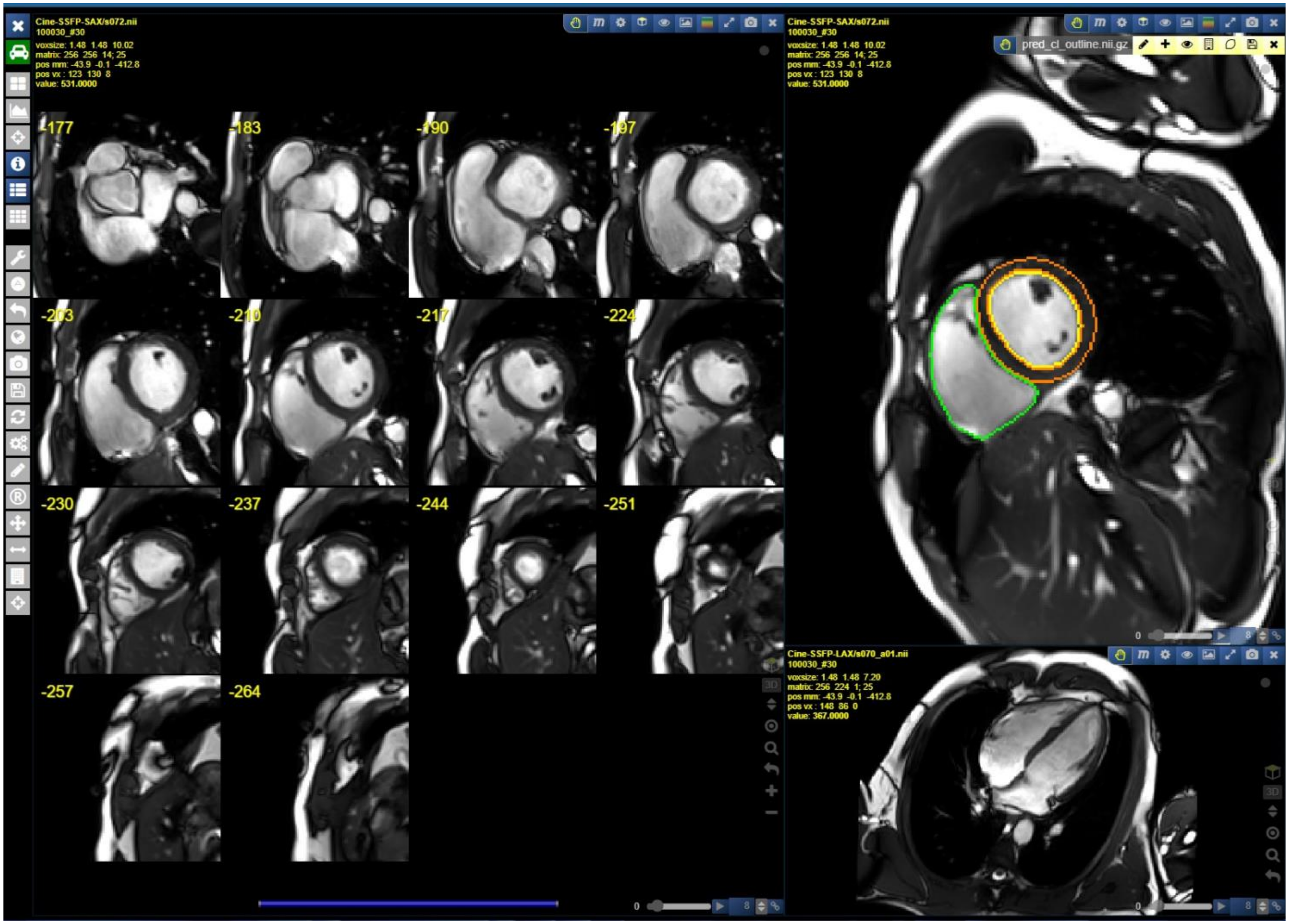
